# Genetic Variants Associated with Placental Abnormalities in Stillbirths

**DOI:** 10.64898/2026.10.19.26365933

**Authors:** Tsegaselassie Workalemahu, Thomas J. Nicholas, Aaron R. Quinlan, Xiaoxu Yang, Laurie B. Griffin, Susannah Leisher, Nathan R. Blue, Anya L. Arthurs, Jessica M. Page, D. Ware Branch, Nicola J. Camp, Lynn B. Jorde, Claire T. Roberts, Jessica L. Giordano, Ronald Wapner, John Kingdom, Karen J. Gibbins, Mana M. Parast, Uma M Reddy, Robert M. Silver

## Abstract

**Background:** Placental insufficiency (PI), characterized by impaired placental function, is a leading cause of stillbirth. Although rare coding variants are known to cause a subset of stillbirths, their contribution to PI is poorly understood.

**Methods:** We conducted a secondary analysis of stillbirth cases (N=292) of the Stillbirth Collaborative Research Network study with exome sequencing available on postmortem placental or fetal samples. PI was defined by standardized placental pathological lesions (e.g., maternal vascular malperfusion) and prespecified clinical criteria for placental dysfunction. Candidate gene-based rare-variant burden analysis was performed using the Sequence Kernel Association Test-Optimal framework, focusing on evolutionarily constrained developmental and placental-function genes. Primary analysis compared stillbirths with and without placental vascular malperfusion. Similar comparisons by clinical PI and other placental pathological lesions were conducted in secondary analysis.

**Results:** Stillbirths with PI phenotypes demonstrated different patterns of genetic variation than stillbirths without PI. Specifically, we observed nominal differential enrichment of constrained variants in biologically prioritized placental-function genes, regulating extravillous trophoblast invasion and spiral artery remodeling (*CTNND2*, *PHACTR1*, *SLIT1, VEGFA*), placental angiogenesis (*IGF2R*, *TEK*), mitochondrial homeostasis (*OPA1*), and fetal growth (*DLK1*, *GNAS*). Secondary analyses on specific placental pathological lesions suggested recurrent enrichment of genes within similar placental developmental pathways.

**Conclusions:** We observed nominal differential enrichment of constrained developmental and placental-function genes in stillbirths with PI compared to stillbirths without PI. The observed genetic signals across PI phenotypes suggest shared biological pathways underlie placental dysfunction. Our findings advance genotype-phenotype characterization and help prioritize candidate pathways for larger collaborative genomic and functional studies to improve stillbirth risk stratification and prevention.

## 1. Introduction

Stillbirth accounts for substantial proportion of perinatal mortality worldwide, with placental insufficiency (PI) representing the most common potentially preventable cause of death in high-resource settings.^1,2^ Broadly defined as poor placental function, PI underlies common adverse pregnancy outcomes that are associated with stillbirth, which include fetal growth restriction and preeclampsia.^3^ These adverse pregnancy outcomes affect at least 10% of pregnancies in the United States.^4^ Placental pathological lesions, characterized by maternal and fetal vascular malperfusion, chronic inflammation, and/or impaired placental development, are frequently identified in stillbirths and provide important mechanistic insights into the pathogenesis of PI.^5^

PI is hypothesized to arise from dysfunction in multiple heterogeneous pathways of critical placental development (e.g., vascular remodeling, trophoblast signaling).^6^ While PI often occurs in the setting of known maternal vascular comorbidities, it also occurs in the absence of these conditions, which suggests possible underlying genetic contribution. Genetic factors directly account for a subset of stillbirths,^6^ but the genetic basis of PI remains largely unknown. Identifying important genetic contributors to PI may support the development of genetic biomarkers of PI, which in turn holds promise for improving diagnosis, estimating recurrence risk, informing prevention strategies, and enabling precision care to prevent adverse pregnancy outcomes.^7^ These findings may also help identify novel therapeutic targets to improve placental function.

Evolutionary constraint metrics characterize genes under strong purifying selection and have been used to prioritize dosage-sensitive genes underlying severe developmental disorders including those affecting fetal development.^8–10^ In the multicenter Stillbirth Collaborative Research Network (SCRN) study, exome sequencing analysis identified likely monogenic diagnoses in 8-10% of unexplained stillbirths.^6^ The study also demonstrated an enrichment of loss-of-function variants in evolutionarily constrained genes, underscoring the importance of constrained genes for severe developmental phenotypes, including stillbirth. In a separate analysis of the same SCRN cohort, we previously demonstrated that pathogenic copy number variants (CNVs), as well as CNVs of uncertain significance, were associated with PI, implicating structural genomic variation in placental dysfunction.^11^

Building on previous studies and given that stillbirth represents an extreme developmental phenotype subject to strong purifying selection,^9,10^ we posit that gene constraint-informed rare variant analyses would elucidate biologically meaningful genes underlying PI. Specifically, we hypothesized that stillbirths with PI are differentially enriched for rare coding variants in evolutionarily constrained genes involved in placental development, compared with stillbirths without PI. Therefore, we performed constraint-informed candidate gene-based rare variant burden analyses comparing stillbirths with and without histopathologically and/or clinically defined PI.

## 2. Methods

### 2.1. Study Design and Population

The present study is a secondary analysis among stillbirth cases of the SCRN study, a multi-site case-control study designed to classify and investigate the causes of stillbirth in the United States.^12,13^ Stillbirth was defined as fetal death at ≥20 weeks’ gestation. The study included racially/ethnically diverse participants from five catchment areas defined *a priori* by state or county boundaries composed of 59 community and academic hospitals. These areas included Brown University, Rhode Island; Emory University, Georgia; University of Texas Medical Branch at Galveston, Texas; University of Texas Health Sciences Center at San Antonio, Texas; and University of Utah Health Sciences Center, Utah. The present study was approved by the IRB of the University of Utah (IRB application no. 127960; date: March 26, 2020).

### 2.2. Placental Insufficiency Defined by Histopathological and Clinical Data

Study participants underwent a standardized maternal interview, medical record abstraction, biospecimen collection, and postmortem examinations of the fetus and placenta. Postmortem examinations and placental histologic examinations were performed by perinatal pathologists who underwent centralized training using a standardized format.^12,13^ A consensus-determined protocol was implemented to define placental pathological lesion groupings as any vascular malperfusion (maternal vascular and/or fetal vascular malperfusion), any inflammatory (maternal and/or fetal chronic inflammatory), and immune/idiopathic lesions.^5^ The overarching (e.g., acute villitis, chronic histiocytic intervillositis, etc.) were included in the categories of the pathologic and have been previously published in detail.^11,13^ The present study evaluated any vascular malperfusion, the most strongly associated category of placental pathological lesion in stillbirth,^5,14^ as the primary outcome.

Given there is uncertainty about placental lesions, grading/staging, and reproducibility,^15^ we additionally evaluated clinical PI as a secondary outcome. Clinical PI was a composite outcome with evidence for any of the following: small-for-gestational age baby (birthweight < 3rd percentile), abnormal umbilical artery Doppler (absent or reverse end diastolic velocities), oligohydramnios (AFI < 5 cm with intact membranes), and/or severe preeclampsia/preeclampsia before 37 weeks’ gestation.^16,17^ In addition, we evaluated a composite outcome defined as stillbirths with either clinical PI or any placental vascular malperfusion. Because PI can occur in both singleton and twin gestations,^18^ we included all stillbirths with adequate exome sequencing (ES) data and without restricting the cohort to unexplained singleton stillbirths (n=296), corresponding to the final analytical cohort reported by Stanely *et al*.^6^ However, after excluding those without standardized placental histopathology data, only singleton stillbirths with adequate ES data and postmortem examinations of the fetus and placenta (n=253) were available for our primary outcome analysis (**Fig S1**).

### 2.3. DNA Extraction, Genotyping and Variant Annotation

Biospecimens collected as part of the study protocol included placental tissue, stored/frozen fetal liver, fetal muscle, or cord blood. Sizes of placental biopsies varied, but they were as large as 1 cm^3^ and were stored at – 20°C from 2-5 years prior to immediate DNA extraction. DNA was extracted from stored placenta, cord blood, frozen muscle or liver specimens using Puregene, Qiagen Systems as detailed previously.^6^ ES of all samples was performed at the Institute for Genomic Medicine at Columbia University. According to standard protocols of bioinformatics pipeline,^6^ exomes were captured using either the IDT xGen Exome Research Panel V1.0 (Integrated DNA Technologies, Coralville, IA, USA), the Agilent All Exon (65 MB; Agilent Technologies, Santa Clara, CA, USA) kit or the Nimblegen SeqCap EZ (version 2·0 or 3·0) Exome Enrichment kit (Roche NimbleGen, Madison, WI, USA) and sequenced according to standard protocols on Illumina’s (Illumina, San Diego, CA, USA) NovaSeq 6000 and HiSeq2500 platforms. ES reads were aligned to the reference genome (Genome Reference Consortium build 37, human genome 19). Variant and genotype calling was previously performed using Genome Analysis Toolkit (GATK; Broad Institute, Boston, MA, USA) Best Practices v3.6. Joint-genotype calling was performed using the raw ES data to generate a unified variant call set using standardized institutional pipeline the University of Utah Center for Genomic Discovery (UCGD). *Somalier*, a genotype-based genetic ancestry/family validation tool, was used to confirm familial relationships in these unrelated probands of stillbirths and ancestries based on the self-reported demographic data.^19^ Left-alignment, variant normalization and call set across all samples, and quality control were completed.

Variants were annotated using Variant Effect Predictor. Previously, ES analysis of stillbirths in the same study analyzed 18,653 genes stratified by “intolerance” to functional variation and found molecular diagnoses in 13.4% (36/268 stillbirths).^6^ Aligned with the previous findings, we posit that genes under strong purifying selection are depleted for protein-truncating variation in the general population; therefore, analyses based on constraint metrics enrich for genes that are dosage-sensitive or loss-of-function (LoF)-intolerant to improve discovery of rare-disease genes.^10^ Thus, we annotated variants using multiple genomic constraint metrics, which included Loss-of-function Observed/Expected Upper bound Fraction (LOEUF) and probability of loss-of-function intolerance (pLI), Constrained Coding Region (CCR) percentiles, and Gnocchi scores. Specifically, LOEUF and pLI measure constraint against predicted LoF variants, where lower LOEUF and higher pLI values indicate greater intolerance to LoF variation.^10^ CCRs are genomic regions within protein-coding sequences that show significant depletion of variation in the general population, where higher CCR scores indicate greater constraint against genetic variation.^9^ Gnocchi is a regional constraint metric that quantifies depletion of rare genetic variation relative to expectation, and higher Gnocchi scores indicate greater intolerance to genetic variation.^8^ In the present study, gene-based rare-variant burden analyses were constraint-informed, i.e., evaluating variants with pLI ≥ 0.9, LOEUF ≤ 0.6, CCR ≥ 95% or GNOCCHI ≥ 4.^8,9^ Lastly, variants were annotated by ClinVar disease associations and curated stillbirth-related gene lists.

### 2.4. Candidate Gene Selection

While PI is likely biologically and genetically heterogeneous, with maternal and fetal-placental underpinnings,^3^ we conducted candidate gene-based analysis evaluating rare variants in functionally relevant genes (N=159; **Table S2**). These included core genes with previously recognized roles in placental development, categorized as: 1) core trophoblast lineage and placental development,^20^ 2) extravillous trophoblast invasion and spiral artery remodeling,^21^ 3) placental angiogenesis and vascular function,^22,23^ 4) Placental/fetal growth,^24,25^ 5) placental mitochondrial/stress-response,^26^ and 6) placenta-specific microRNA/ncRNA systems.^23,27^ Accordingly, gene-based rare variant burden analyses were hypothesis-based, interpreting results based on these genes with relevant etiology.

### 2.5. Statistical analysis

We conducted rare-variant association testing using Sequence Kernel Association Test-Optimal (SKAT-O).^28^ Briefly, SKAT-O tests whether the aggregate genotype distribution differs between groups (testing the *null* hypothesis that the aggregate distribution of qualifying rare variants within a gene is the same between groups). SKAT-O with small-sample adjustment was applied to combine burden and variance-component testing and to control Type I error in small samples.^29,30^ In SKAT-O models, genetic ancestry (broad differences among African/Admixed-American or Native American/European ancestry groups) and tissue source (placenta vs other source) were adjusted as covariates. For each gene, SKAT-O calculated Rank, a genotype matrix rank representing the number of truly independent variants contributing to gene burden after accounting for identical patterns/correlation and total minor allele count (MAC) across all qualifying variants. P-value is estimated from the corresponding *null* distribution of a variant-component score statistic.^28^ Genes with genotype-matrix Rank ≥ 3 and MAC ≥ 5, corresponding to gene-level tests supported by multiple independent genotype dimensions and at least five observed minor alleles, were summarized.^31^ Genes with nominal statistical significance (SKAT-O P-value < 0.05) were prioritized for biological interpretation. Given the uncertain functional consequences and potentially opposing effects of variants, we interpreted SKAT-O associations without inferring direction of effect from carrier-count differences.

Because confined placental mosaicism is associated with placental dysfunction and adverse pregnancy outcomes,^32^ we evaluated whether the SKAT-O prioritized genes were previously implicated in human mosaic disorders. The observed gene set was compared with MosaicBase,^33^ a curated database of non-cancer mosaic variants. Gene labels from 19,909 human genes were randomly permuted 10,000 times and the overlap with MosaicBase genes was recalculated for each permutation to generate an empirical *null* distribution. An empirical permutation *P-value* was determined as the proportion of permutations with overlap greater than or equal to the observed overlap. Analyses and data visualization were performed using R version 4.5.0.

## 3. Results

Among 295 stillbirths with ES data, 292 (99 with PI and 193 without PI) had complete covariate data and were included in our secondary clinical PI analysis (**Table 1** and **Fig S1**). Of these, 253 (174 with any placental vascular malperfusion and 79 without any placental vascular malperfusion) had standardized placental histopathology data available for our primary analysis. Among 99 stillbirths with clinical PI, 74 stillbirths also had placental vascular malperfusion. Maternal age, history of previous stillbirth, baby sex distributions were similar in stillbirths with any of PI phenotypes, compared with those without PI phenotypes. Stillbirths with PI phenotypes were characterized by earlier gestational age at delivery, lower birthweight, higher frequencies of small-for-gestational-age baby, and greater prevalence of preeclampsia than their counterparts. Although overall genetic ancestry distributions were not statistically significantly different by PI phenotypes, placental vascular malperfusion occurred more frequently among stillbirths of African and Admixed-American/Native-American ancestry than among those of European ancestry.

**Table 1.** Clinical characteristics of stillbirths by placental histopathology and placental insufficiency phenotypes.

|  | Any Placental Vascular Malperfusion (N=253) |  |  | Clinical Placental Insufficiency (N=292) |  |  | Clinical Placental Insufficiency and/or Any Placental Vascular Malperfusion (N=292) |  |  |
| --- | --- | --- | --- | --- | --- | --- | --- | --- | --- |
| Characteristics<sup>1</sup> | Yes | No | P-value<sup>2</sup> | Yes | No | P-value<sup>2</sup> | Yes | No | P-value<sup>2</sup> |
| N | 174 | 79 |  | 99 | 193 |  | 199 | 93 |  |
| Maternal age at delivery, yrs | 27.2 ± 6.7 | 27.5 ± 6.6 | 0.99 | 28.1 ± 6.7 | 27.0 ± 6.6 | 0.29 | 27.3 ± 6.7 | 27.6 ± 6.4 | 0.88 |
| Gestational age at delivery, wks | 28.3 ± 6.5 | 29.7 ± 7.1 | 0.15 | 26.9 ± 5.6 | 29.0 ± 7.0 | 0.02 | 28.1 ± 6.4 | 28.6 ± 7.0 | 0.63 |
| Birthweight, g | 1299 ± 1143 | 1688 ± 1272 | 0.03 | 845 ± 736 | 1606 ± 1265 | <0.001 | 1238 ± 1112 | 1551 ± 1250 | 0.03 |
| Small-for-gestational age | 72 (41.4) | 18 (22.8) | 0.005 | 87 (87.9) | 14 (7.3) | <0.001 | 95 (47.7) | 6 (6.5) | <0.001 |
| Male baby | 93 (53.8) | 38 (48.1) | 0.42 | 52 (52.5) | 96 (50.3) | 0.80 | 102 (51.5) | 46 (50.0) | 0.90 |
| Gestational Hypertension | 18 (10.3) | 8 (10.3) | 0.99 | 16 (16.2) | 14 (7.3) | 0.03 | 24 (12.1) | 6 (6.6) | 0.21 |
| Preeclampsia | 20 (12.0) | 2 (2.5) | 0.02 | 22 (23.2) | 2 (1.1) | <0.001 | 24 (12.6) | 0 (0.0) | <0.001 |
| Nulliparity | 80 (46.0) | 31 (39.7) | 0.41 | 47 (47.5) | 78 (40.6) | 0.32 | 94 (47.2) | 31 (33.7) | 0.03 |
| Previous stillbirth | 14 (8.0) | 5 (6.2) | 0.59 | 7 (7.0) | 16 (8.2) | 0.64 | 15 (7.5) | 8 (8.5) | 0.99 |
| Genetic Ancestry |  |  | 0.49 |  |  | 0.25 |  |  | 0.81 |
| European | 59 (33.9) | 34 (43.0) |  | 36 (36.4) | 70 (36.3) |  | 72 (36.2) | 34 (36.6) |  |
| Admixed-American/Native-American | 68 (39.1) | 26 (32.9) |  | 32 (32.3) | 76 (39.4) |  | 71 (35.7) | 37 (39.8) |  |
| African | 42 (24.1) | 16 (20.3) |  | 25 (25.3) | 43 (22.3) |  | 48 (24.1) | 20 (21.5) |  |

<sup>1</sup> Values are mean ± standard deviation for continuous variables and N (%) for categorical variables
<sup>2</sup> P-values were calculated using the Wilcoxon rank-sum test for continuous variables and Fisher's exact test for categorical variables.

Overall, 151,386 rare variants with non-missing gene annotations representing 19,413 genes, including 16,898 protein-coding genes, were evaluated in gene-based burden analyses. Quantile-quantile plot indicated a moderate inflation factor (lambda=1.3) with some deviations from the expectation, suggesting a limited subset of genes contributed disproportionately to the observed signal (**Fig S2**). **Fig S3** shows Manhattan plots with annotated placental function genes passing the SKAT-O P-value=0.05 significance line indicated in blue, and the Bonferroni-adjusted significance line shown in red, which none exceeded.

We observed nominal differential gene-level rare-variant enrichment in constrained and biologically prioritized placental-function genes across all three phenotypes (**Table 2**). Among stillbirths with placental vascular malperfusion, nominal rare-variant enrichment was observed for *CTNND2* (P-value=0.003), *PHACTR1* (P-value=0.02), *SLIT1* (P-value=0.04), *OPA1* (P-value=0.007), and *IGF2R* (P-value=0.02) genes. Among stillbirths with clinically defined PI, nominal rare-variant enrichment was observed for *VEGFA* (P-value=0.02), *SLIT1* (P-value=0.03), *TEK* (P-value=0.01), and *DLK1* (P-value=0.04) genes. Enrichment analyses by combined placental vascular malperfusion/clinical PI phenotype recapitulated recurrent nominally significant signals in *PHACTR1* (P-value=0.03) and *OPA1* (P-value=0.004) genes and novel signals in *GNAS* (P-value=0.02) gene. The associated genes mapped predominantly to pathways involving trophoblast invasion/spiral artery remodeling, placental angiogenesis, mitochondrial/stress response, and placental/fetal growth, with recurrent enrichment of variants in *PHACTR1*, *OPA1*, *SLIT1* genes across PI phenotypes (**Fig S4**). None of these nominal associations remained significant after correction for multiple testing.

**Table 2.** Rare variant burden in constrained placental function candidate genes across placental insufficiency phenotypes in stillbirths.

| Gene | Chr | Rare Variant Count<sup>1</sup> | P-value<sup>2</sup> | Nomenclature | Placental Biological Pathway |
| --- | --- | --- | --- | --- | --- |
| **Any Placental Vascular Malperfusion** |  |  |  |  |  |
| CTNND2 | 5 | 9 | 0.003 | Catenin delta 2 | Extra villous trophoblast invasion/spiral artery remodeling |
| PHACTR1 | 6 | 11 | 0.015 | Phosphatase and actin regulator 1 |  |
| SLIT1 | 10 | 20 | 0.035 | Slit guidance ligand 1 |  |
| OPA1 | 3 | 7 | 0.007 | OPA1 mitochondrial dynamin like GTPase | Mitochondrial/stress-response |
| IGF2R | 6 | 20 | 0.023 | Insulin like growth factor 2 receptor | Angiogenesis |
| **Clinical Placental Insufficiency** |  |  |  |  |  |
| VEGFA | 6 | 4 | 0.016 | Vascular endothelial growth factor A | Extra villous trophoblast invasion/spiral artery remodeling |
| SLIT1 | 10 | 20 | 0.029 | Slit guidance ligand 1 |  |
| DLK1 | 14 | 6 | 0.041 | Delta like non-canonical Notch ligand 1 | Placental/fetal growth |
| TEK | 9 | 6 | 0.010 | TEK receptor tyrosine kinase | Angiogenesis |
| **Clinical Placental Insufficiency and/or Any Placental Vascular Malperfusion** |  |  |  |  |  |
| PHACTR1 | 6 | 11 | 0.029 | Phosphatase and actin regulator 1 | Extra villous trophoblast invasion/spiral artery remodeling |
| GNAS | 20 | 12 | 0.021 | GNAS complex locus | Placental/fetal growth |
| OPA1 | 3 | 7 | 0.004 | OPA1 mitochondrial dynamin like GTPase | Mitochondrial/stress-response |

<sup>1</sup> Number of qualifying rare variants contributing to the gene-based burden test.
<sup>2</sup> Gene-level P-value from a Chi-squared distribution. SKAT-O gene-based burden test, testing the aggregate association of qualifying rare variants within each gene with the phenotype after adjustment for ancestry and tissue source.

Secondary analyses of individual placental pathological lesion phenotypes identified additional nominal signals and signals that recapitulated the primary findings (**Table S1**). Specifically, recurring signals in genes *CTNND2*, *SLIT1*, *TNS1*, *DLK1*, and *RICTOR* were observed across multiple placental pathological lesion subtypes. Qualifying rare variants contributing to the gene-level burden signals for all phenotypes are summarized in **Table S3**. Eight of the nine prioritized genes were represented in MosaicBase, significantly exceeding the overlap expected by chance in 10,000 random gene permutations (empirical P-value=0.0007; **Table S4**, **Fig. S5**).

## 4. Discussion

Stillbirths with placental abnormalities demonstrated different patterns of genetic variation than stillbirths without placental abnormalities. There was nominal differential enrichment of variants in constrained placental-function genes with convergent pathways regulating trophoblast invasion, placental vascular development and fetal growth, when comparing stillbirths with and without PI. Although these observations require independent replication and functional validation, the findings highlight multiple developmental mechanisms that underlie PI and may contribute to stillbirth.

The recurrence of genes involved in placental developmental pathways (e.g., trophoblast invasion and spiral artery remodeling) across complementary PI phenotypes (pathological and clinical definitions) in stillbirths is noteworthy. Genomic studies of stillbirth mainly assessed stillbirth as a single, biologically heterogeneous outcome.^6,34^ This approach is useful for identifying genetic causes of death in a subset of stillbirths broadly. However, substantial etiologic heterogeneity is inherent to stillbirth, encompassing PI, fetal structural and genetic disorders, infection, maternal disease, and other mechanisms.^1,35^ The evaluation of clinically and histopathologically defined complementary PI phenotypes in our strategy may have reduced biological heterogeneity and enabled identification of distinct and recurring developmental pathways that can be obscured in analyses of all stillbirths combined.

Trophoblast invasion and spiral-artery remodeling establish the maternal utero-placental circulation, while placental angiogenesis and fetal vascular development determine the capacity of the chorionic placental villi to facilitate feto-maternal oxygen and nutrient exchange to the fetus.^36^ Perturbations in either process can lead to impaired fetal growth and many adverse pregnancy outcomes.^37^ Although clinical and histopathologic manifestations of PI may coexist (e.g., 74 stillbirths with clinical PI also had evidence of placental vascular malperfusion), the recurrence of biological pathways across our complementary PI phenotypes may suggest that shared developmental mechanisms contribute to different placental abnormalities. This interpretation is consistent with the recommendation for standardized criteria to distinguish biologically meaningful placental abnormalities.^14,38^

Previously, genes under strong purifying selection that may be relevant to survival during fetal development have been reported.^6^ Our findings extend this previous finding by showing that evolutionary constraint combined with placental phenotype analyses may help prioritize genes involved specifically in placental development and function relevant to stillbirth. Some of the prioritized genes highlighted in our study (*DLK1*, *PAPPA2*, *MTOR* and *TIMP2*) have also been implicated in placental, fetal-growth, or pregnancy-loss biology in other studies.^39–42^ Therefore, phenotype-guided rare-variant analyses may clarify recurrent developmental mechanisms through which diverse genetic perturbations converge on placental dysfunction. Rare variants in *OPA1*, a mitochondrial/stress-response gene with established haploinsufficiency mechanism, are associated with placental vascular malperfusion and combined clinical PI and placental vascular malperfusion phenotype in our study. However, the contributing variants lacked definitive loss-of-function annotations (**Table S3**), and their potential haploinsufficiency mechanism on disease and effects on mitochondrial function require further investigation.

Our observation involving *DLK1* loci is particularly notable because we have previously reported an inherited 14q32 region overlapping with *DLK1* among stillbirths from a multi-generational family affected by multiple stillbirths.^43^ *DLK1* is a highly conserved gene with increased placental/fetal expression in late gestation.^44,45^ Reduced maternal *DLK1* concentrations are associated with pathologic fetal growth restriction.^45^ Additionally, Page *et al*.^39^ found that low maternal circulating *DLK1* was more frequent among preterm pregnancies with PI, although *DLK1* concentrations were not independently associated with stillbirth. These findings confirm a relationship between the locus and placental/fetal growth biology, although functional studies are required to determine the role of *DLK1* gene variants as a diagnostic markers of PI.

Our study has several limitations. The findings are based on a modest sample size and nominal SKAT-O associations that did not withstand multiple-testing correction. Independent replication and functional validation may clarify the observed findings in this study. Maternal cell contamination testing was not performed in this study, although samples from fetal facing side were obtained and washed during sample collection to get rid of any maternal blood prior to sample storage. Tissue source was included as a covariate in all gene-based models; however, the study was not designed or powered to test tissue-specific genetic enrichment. Although the absence of matched multi-tissue variant calls precluded formal assessment of placenta-restricted associations or confined placental mosaicism, the enrichment of prioritized genes in our study within MosaicBase suggests that pathways implicated in PI overlap with genes known to cause human disease through mosaic variation. Whether postzygotic mosaic mutations contribute to PI and stillbirth warrants investigation using dedicated mosaic variant-calling approaches and matched placental-fetal sequencing. In addition, the absence of parental sequencing limited assessment of variant inheritance patterns and *de novo* variants. The findings from this study do not permit treatment or necessarily improved outcomes in the index stillbirth, however, if confirmed in studies with parental data, it could permit testing of parents to aid in estimating risk of recurrence. If parents learned they were carriers of the implicated gene, they could use this information to guide decisions regarding future reproduction, including use of donor gametes or *in vitro* fertilization with preimplantation genetic testing. Future pregnancies that are determined to carry these variants through diagnostic genetic testing could be candidates for studies of novel treatments to support placental health.

While we evaluated both Clinical PI and placental vascular malperfusion separately and in combination, the phenotypes were complementary but are overlapping phenotypic definitions of placental dysfunction. Placental pathological lesions represent complex biological processes, and although standardized Amsterdam criteria were used for each category of placental pathological lesions,^5^ some degree of lesion misclassification is possible. Our hypothesis-driven analysis focused on biologically curated placental-function genes and may therefore have missed novel genes or pathways contributing to PI. Although we have extended our previous findings for copy number variants associated with PI in stillbirths,^11^ in this ES-based study, we have not evaluated noncoding, structural, and epigenetic mechanisms that likely contribute to PI.

However, several strengths should be acknowledged. We leveraged a large, geographically and ancestrally diverse, well phenotyped stillbirth cohort with standardized placental histopathology and clinical PI data. By integrating evolutionary constraint metrics with biologically curated placental-function genes, and analyzing clinically and histopathologically defined PI, we observed convergent and distinct pathway-level signals across PI phenotypes.

In conclusion, we observed nominal differential enrichment of constrained developmental, placental function genes in stillbirths with placental insufficiency, compared with stillbirths without placental insufficiency. The observed genetic signals across placental insufficiency phenotypes suggest shared biological pathways underlie placental dysfunction. Our findings advance genotype-phenotype characterization and help prioritize candidate pathways for larger collaborative genomic and functional studies to improve stillbirth risk stratification and prevention.

## Data Availability

The stillbirth collaborative research network (SCRN) data are available through the Eunice Kennedy Shriver National Institute of Child Health and Human Development (NICHD) Data and Specimen Hub (DASH), subject to application and approval. Genomic sequencing data are not publicly available because participant consent does not permit unrestricted sharing; access may be considered for qualified investigators upon reasonable request and subject to applicable data-use requirements.

## Acknowledgements

We thank the participants and their families who make this research possible. Genetic sequence alignment and variant calling were performed by the Utah Center for Genetic Discovery Core, part of the Health Sciences Center Cores at the University of Utah. This work utilized resources and support from the Center for High Performance Computing at the University of Utah, partially funded by NIH Shared Instrumentation Grant 1S10OD021644-01A1. The computational resources used were partially funded by the NIH Shared Instrumentation grant 1S10OD021644-01A1. Computational infrastructure support from the Center for High Performance Computing at the University of Utah is gratefully acknowledged.

## Supplementary Figures and Tables

**Fig S1.**
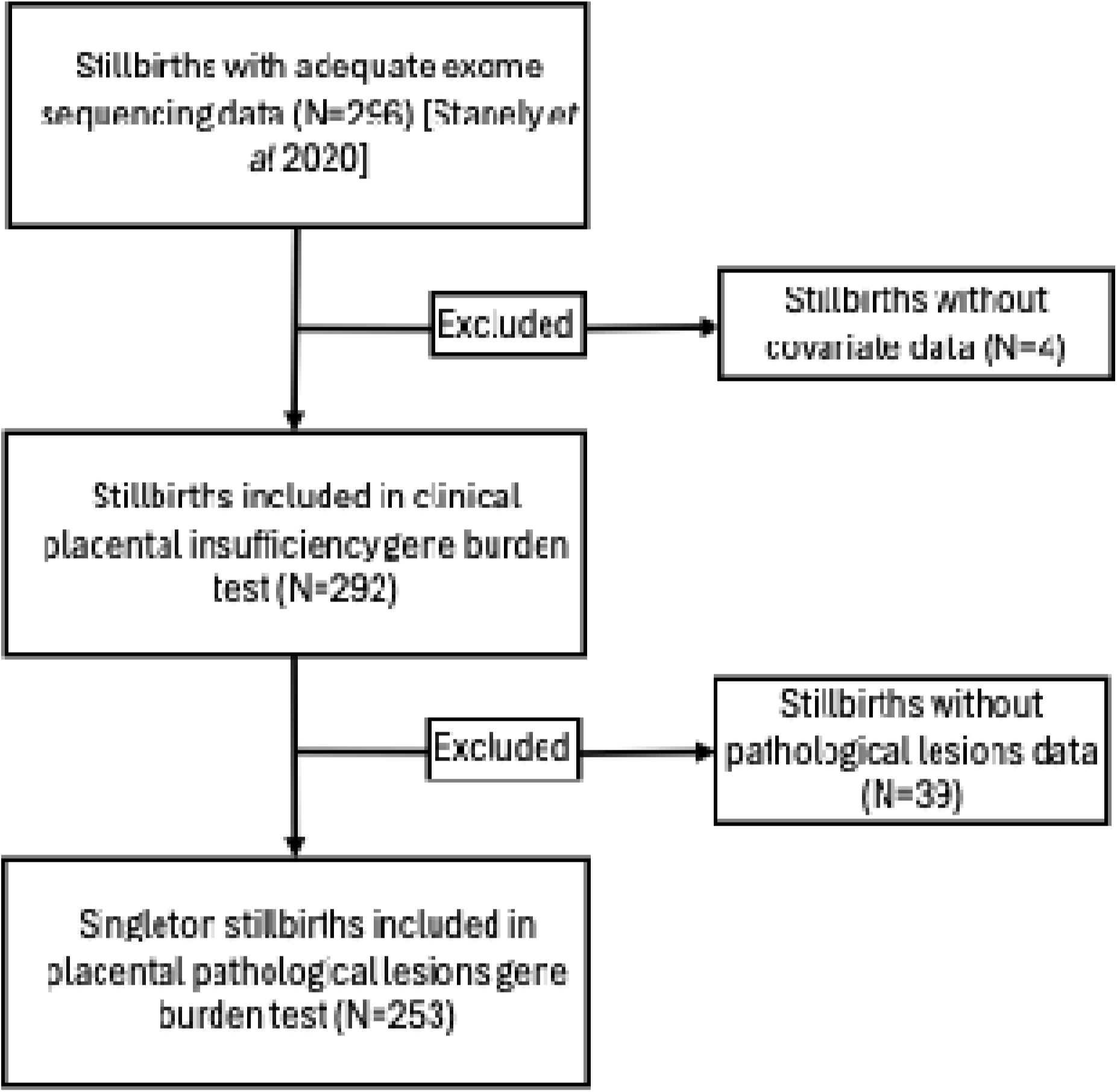
Stillbirth cases included in gene-based rare variant burden analyses

**Fig S2.**
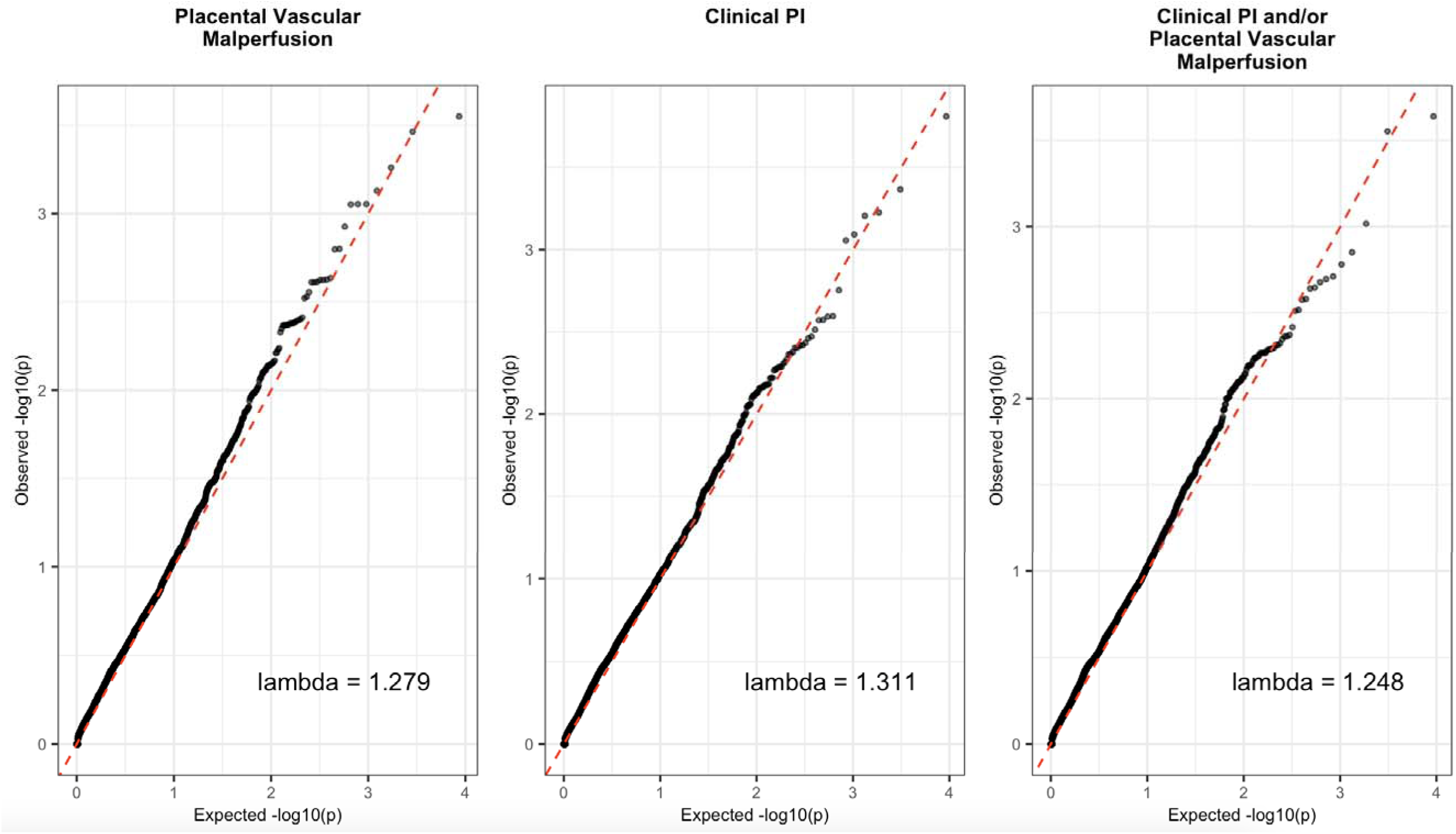
Quantile-quantile plot of constraint-informed gene burden test P-values

**Fig S3.**
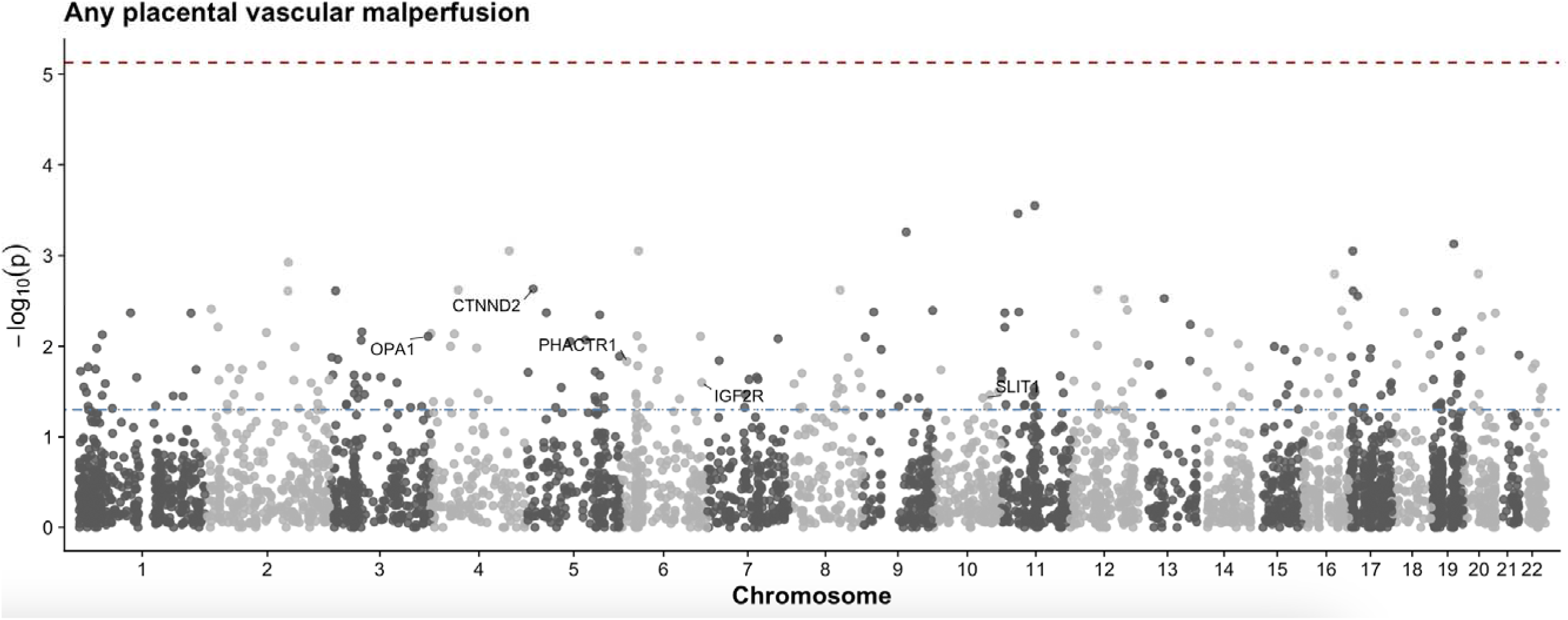
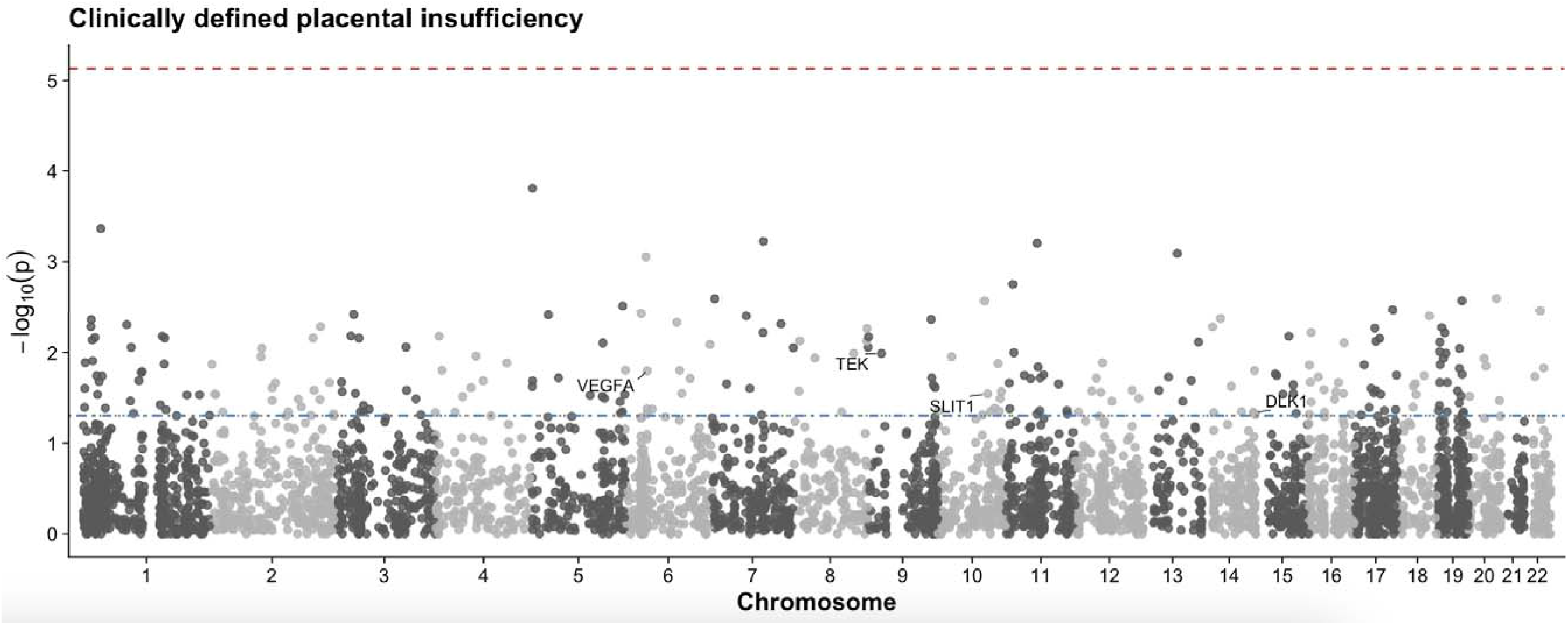
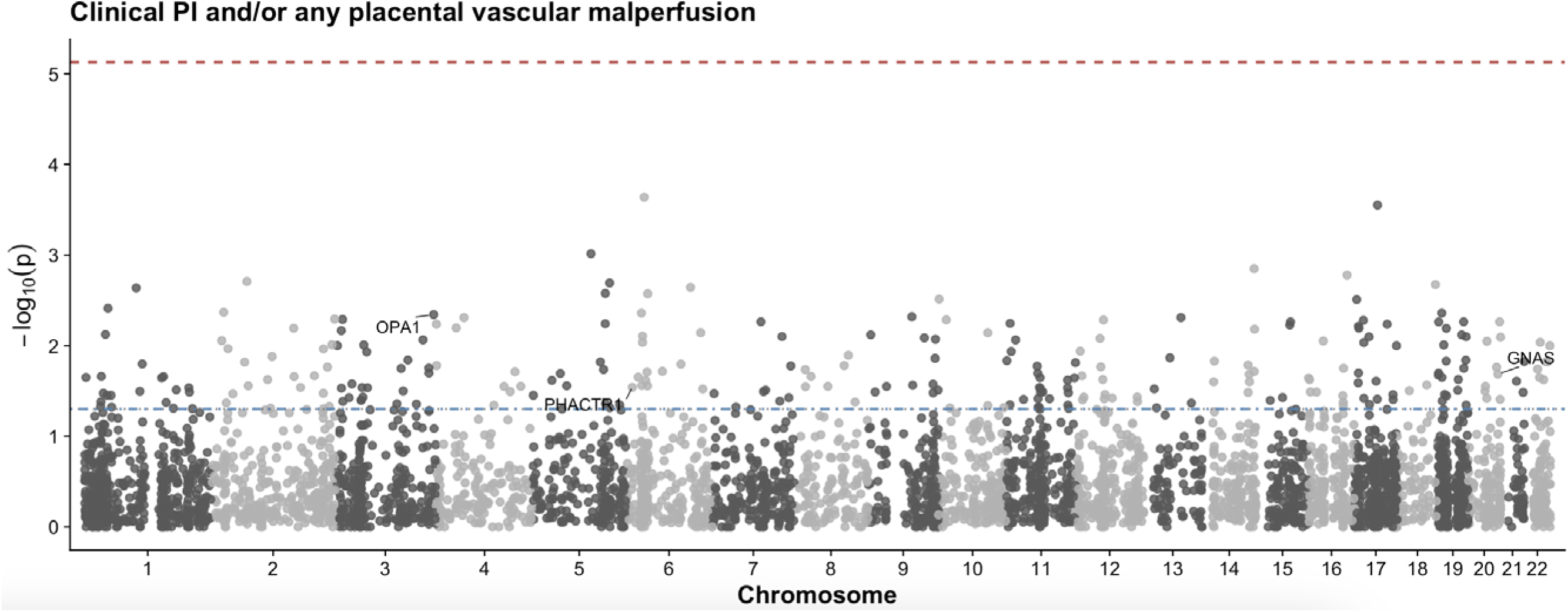
A-C. Manhattan plots with annotated placental function genes. SKAT-O P-value=0.05 significance line is indicated in blue horizontal line, and the Bonferroni-adjusted significance line shown in red horizontal line.

**Fig S4.**
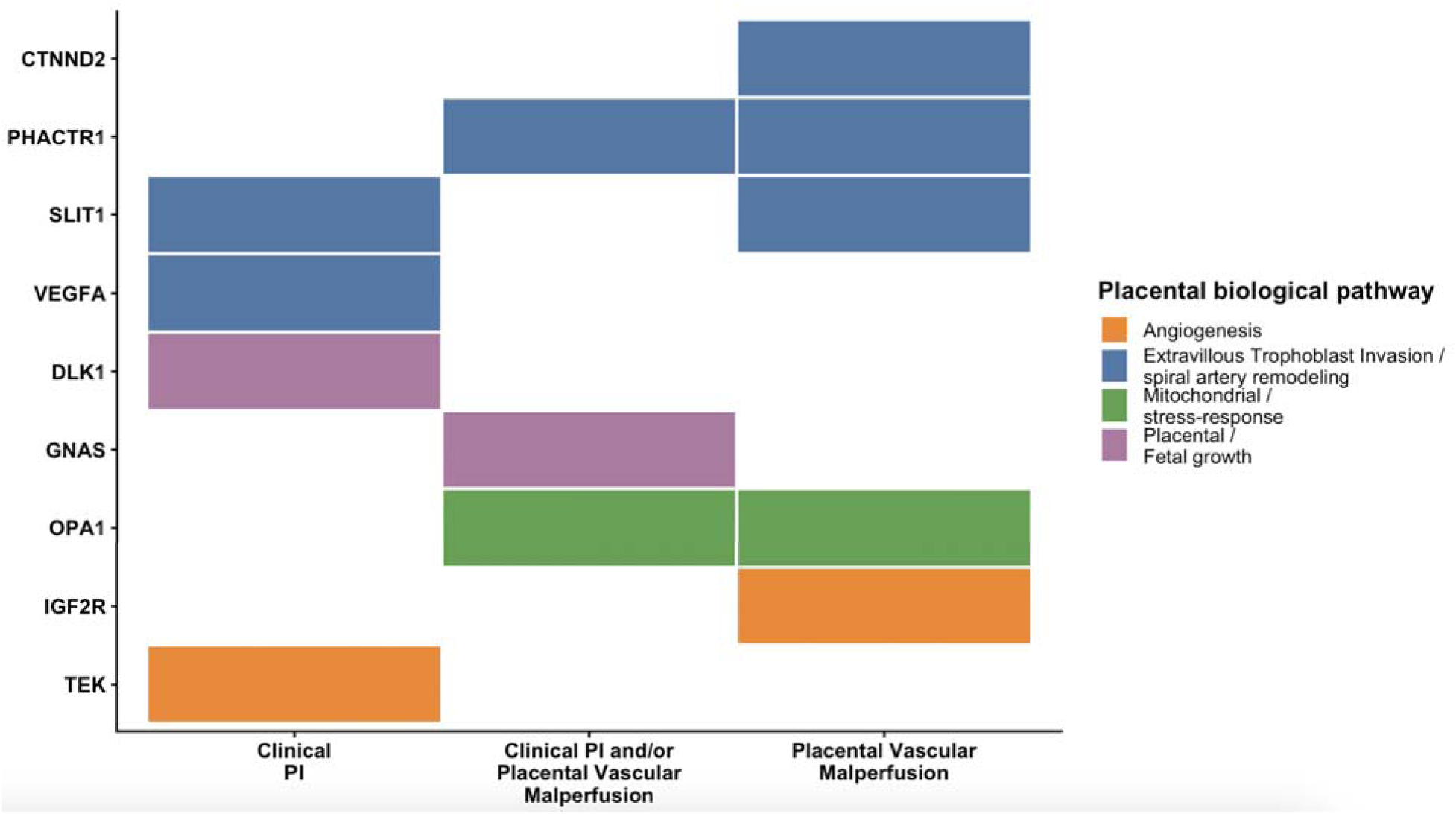
Convergence of biologically prioritized genes across placental insufficiency (PI) phenotypes

**Fig S5.**
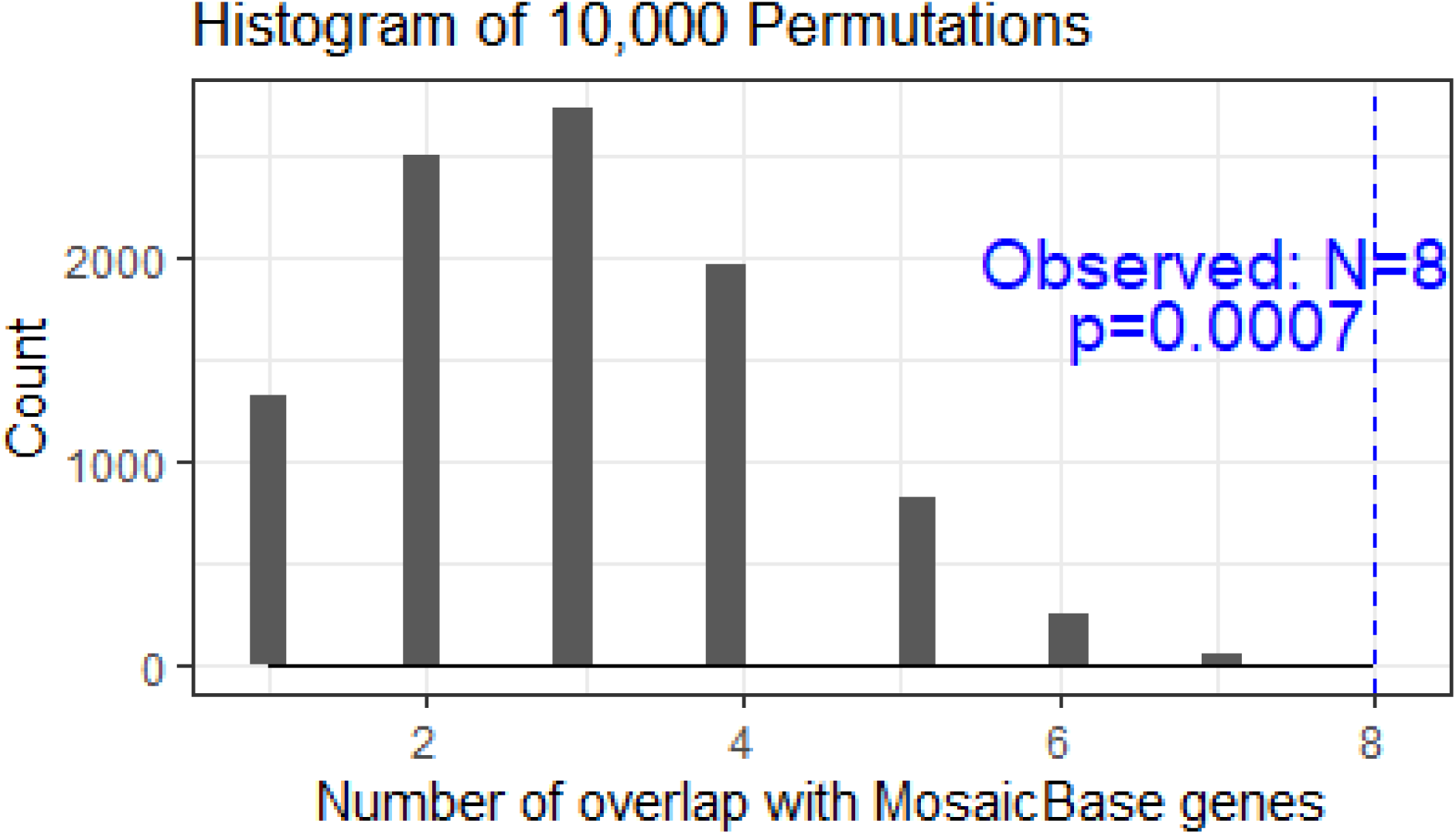
Prioritized genes represented in MosaicBase, significantly exceeding the overlap expected by chance in 10,000 random gene permutations (empirical P-value=0.0007).

**Table S1.**
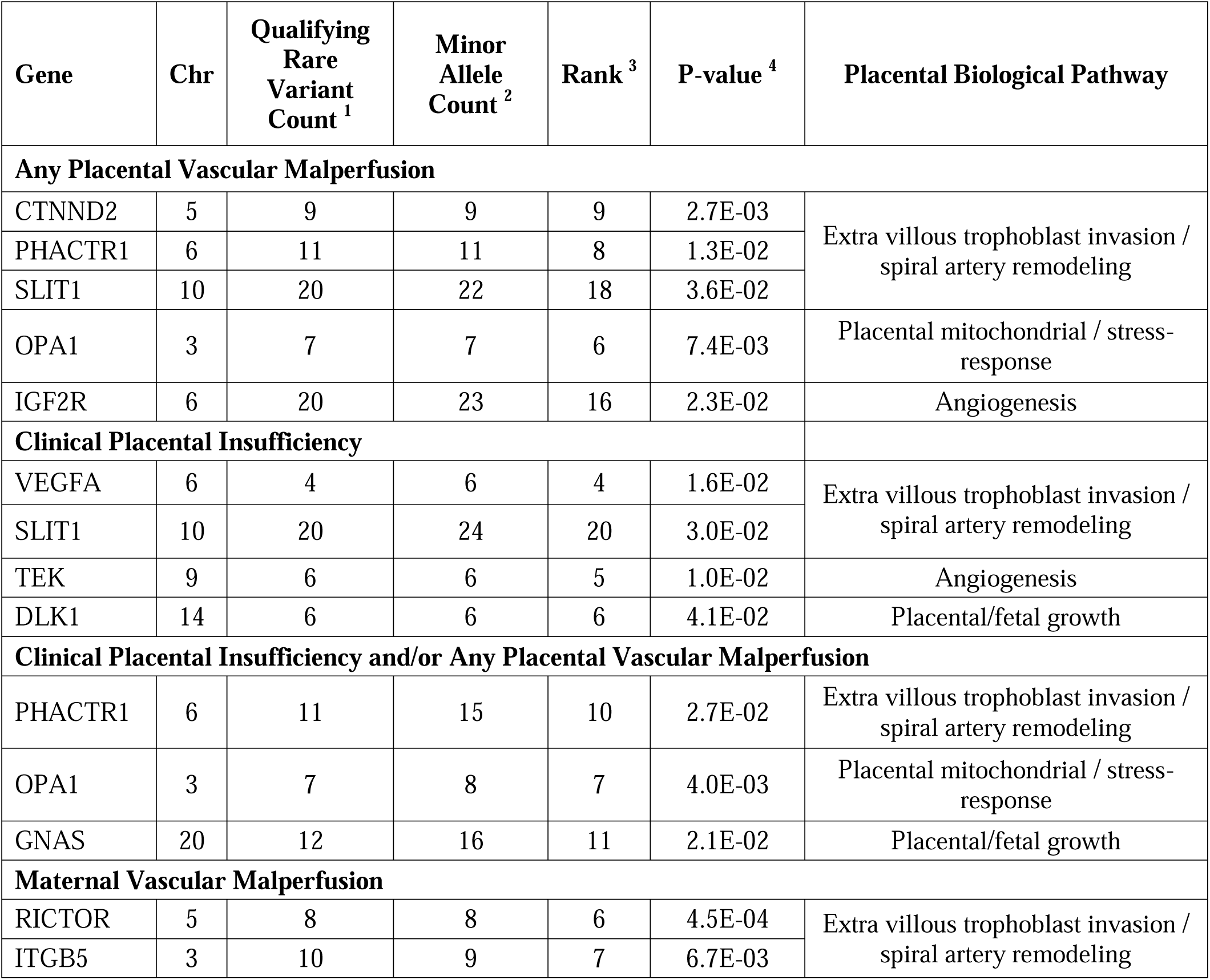
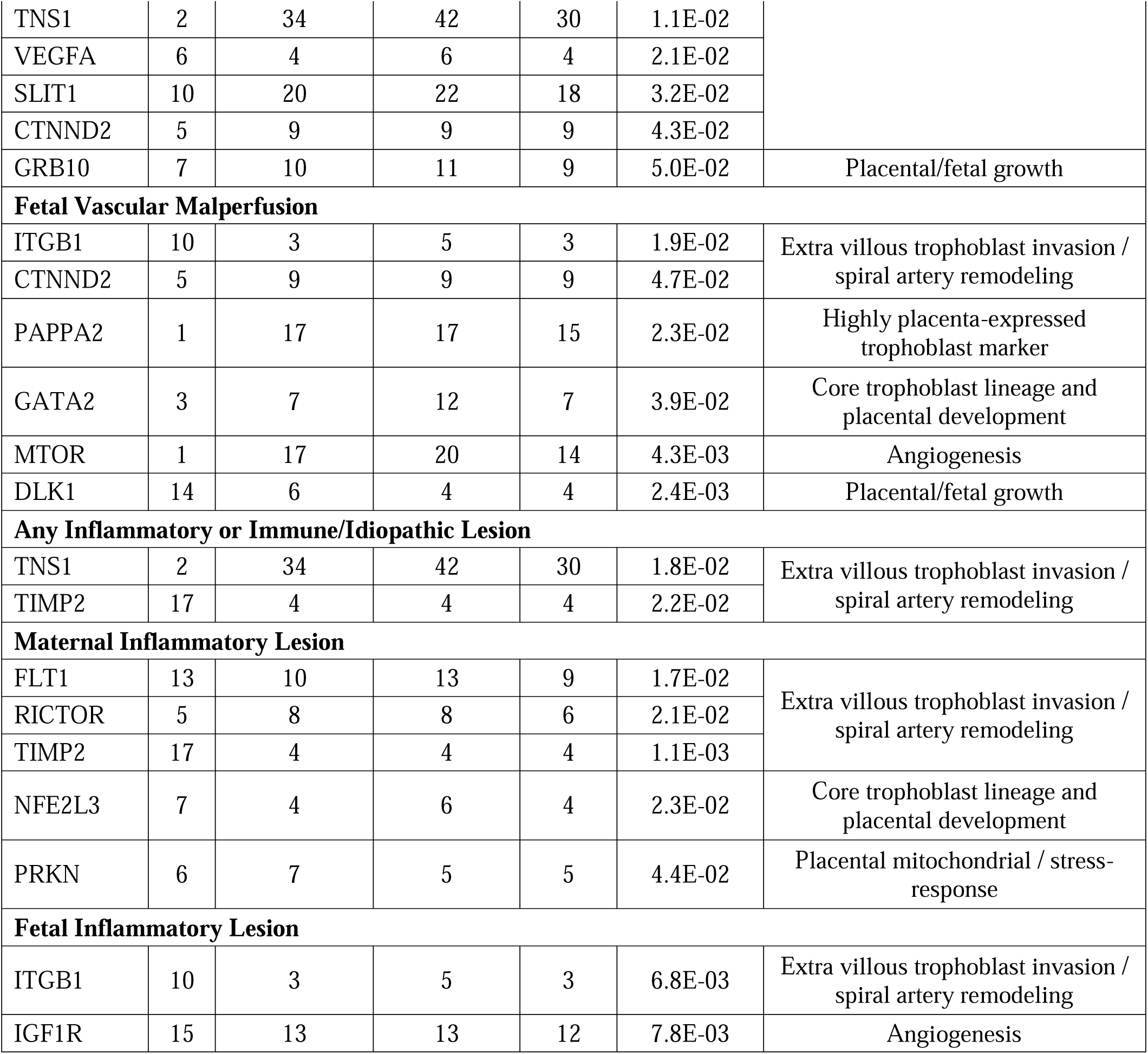
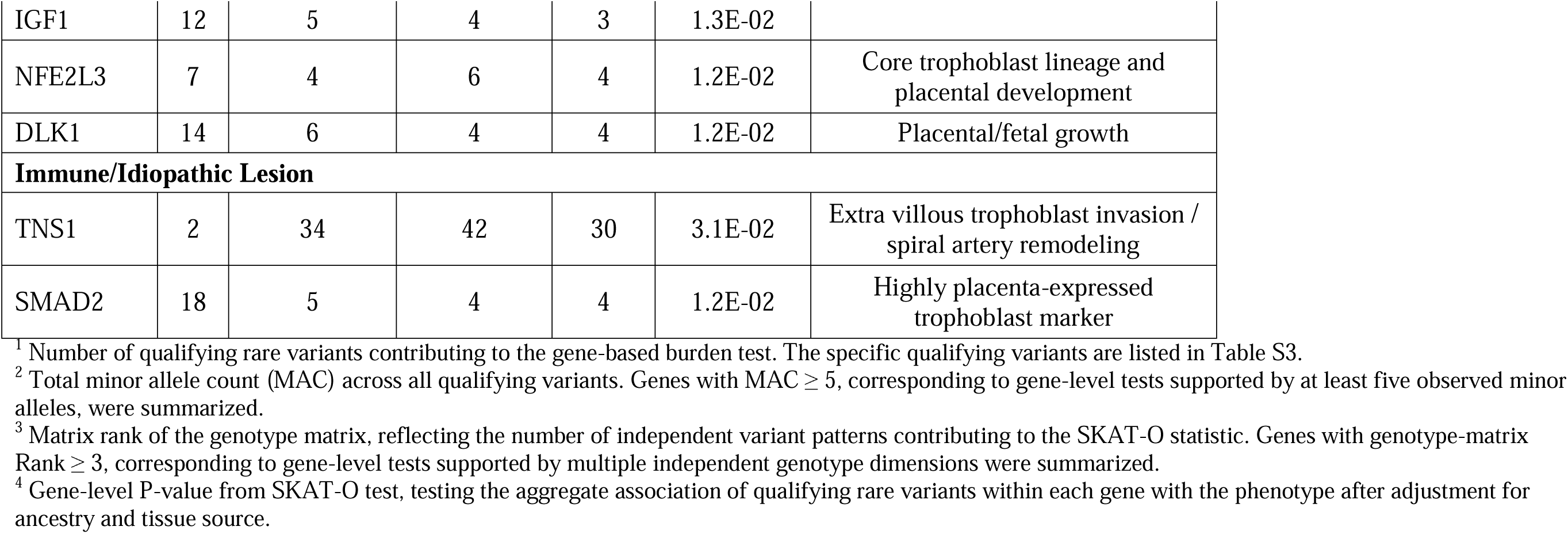
Rare variant burden in constrained placental function candidate genes across placental insufficiency phenotypes, including other pathological lesions in stillbirths.

**Table S2.**
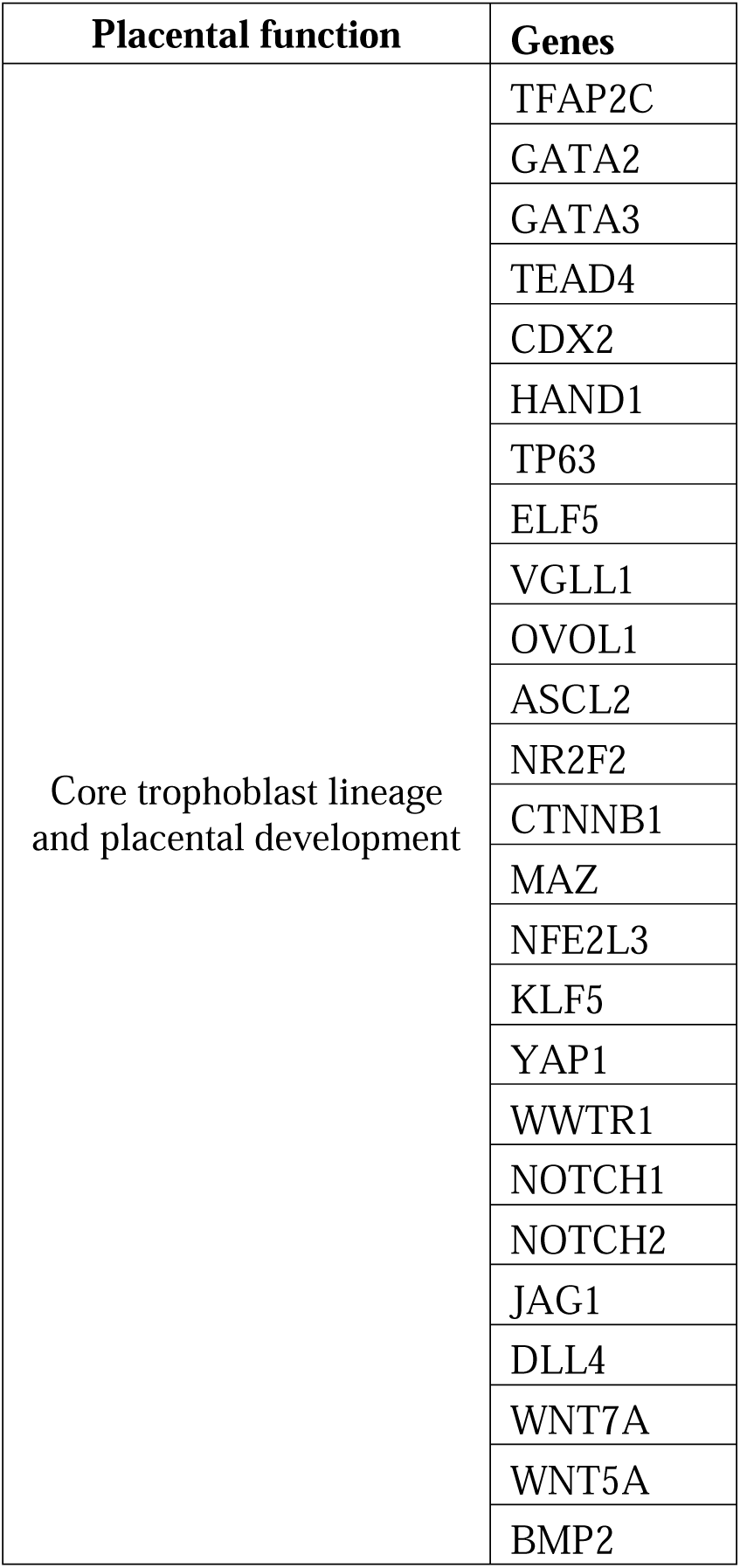
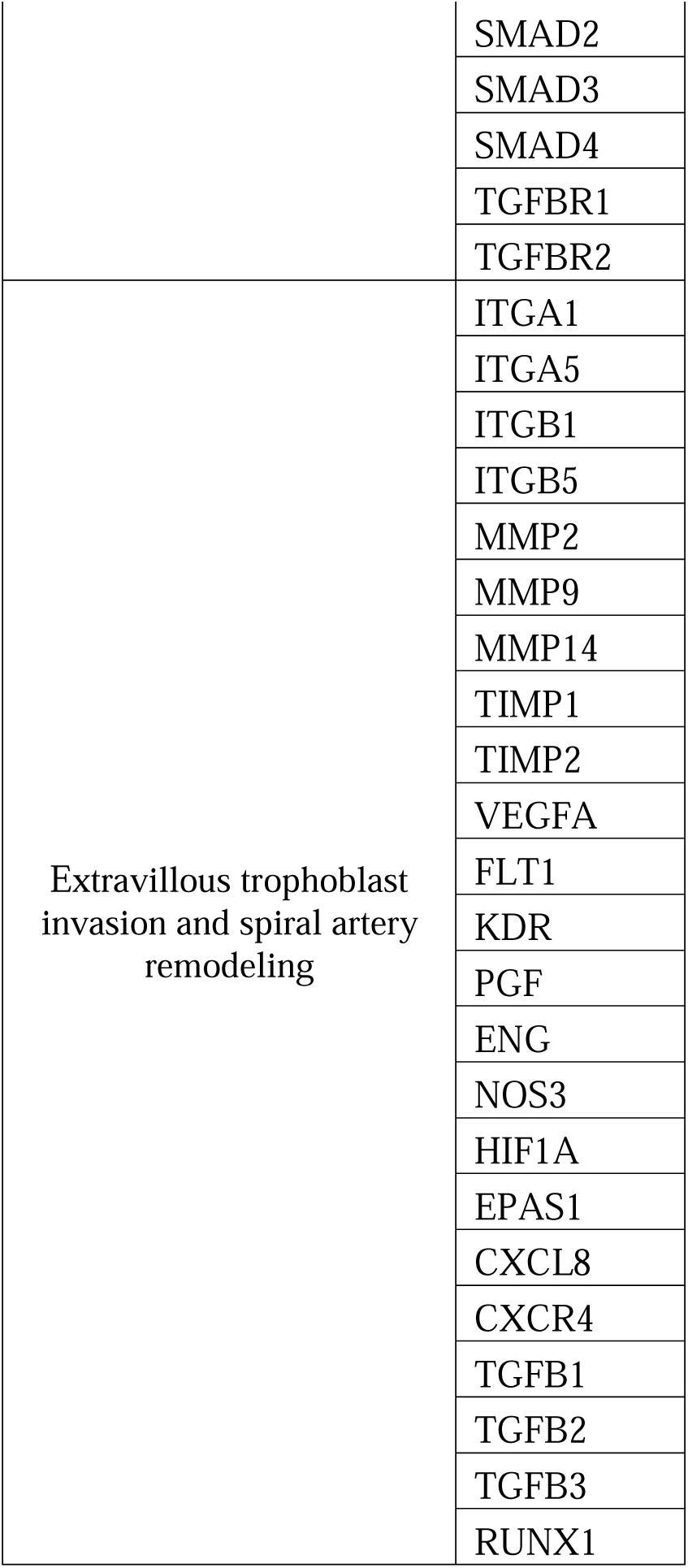
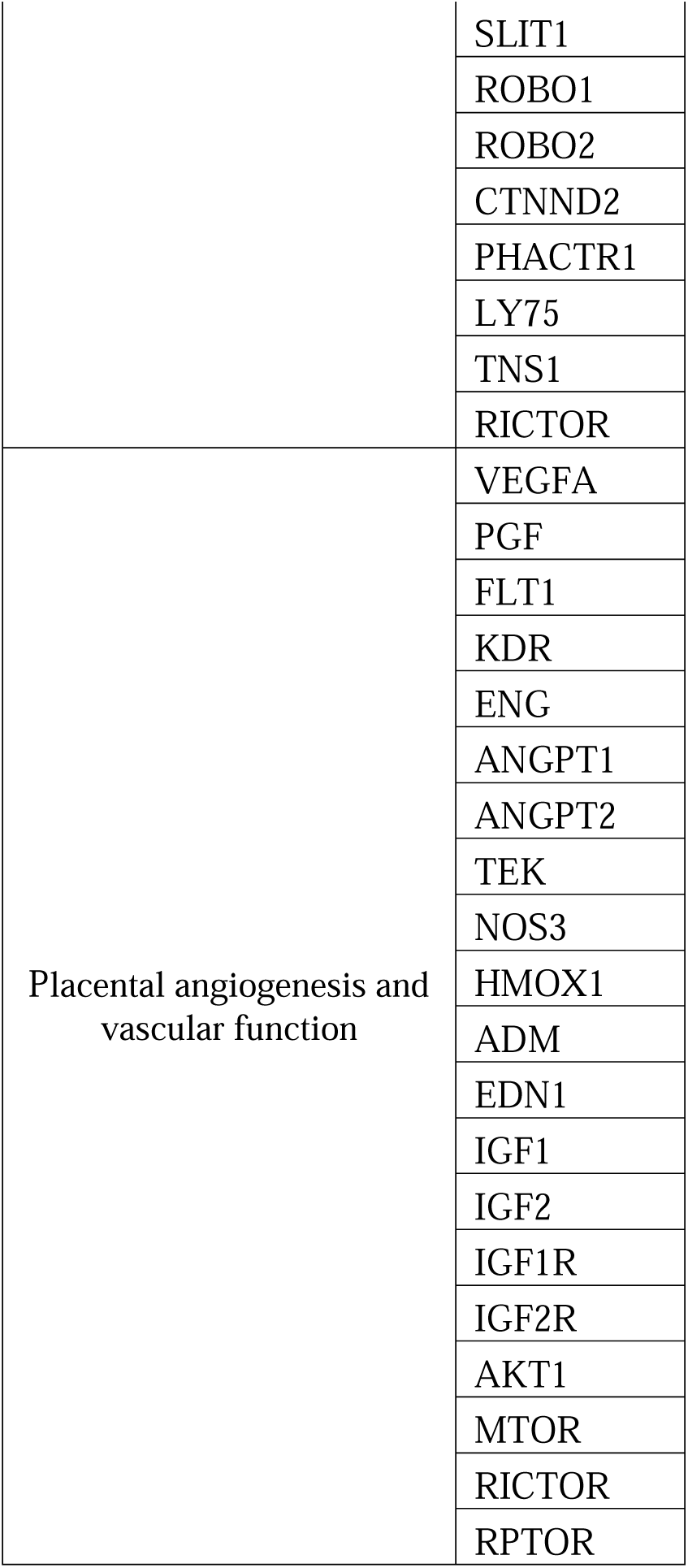
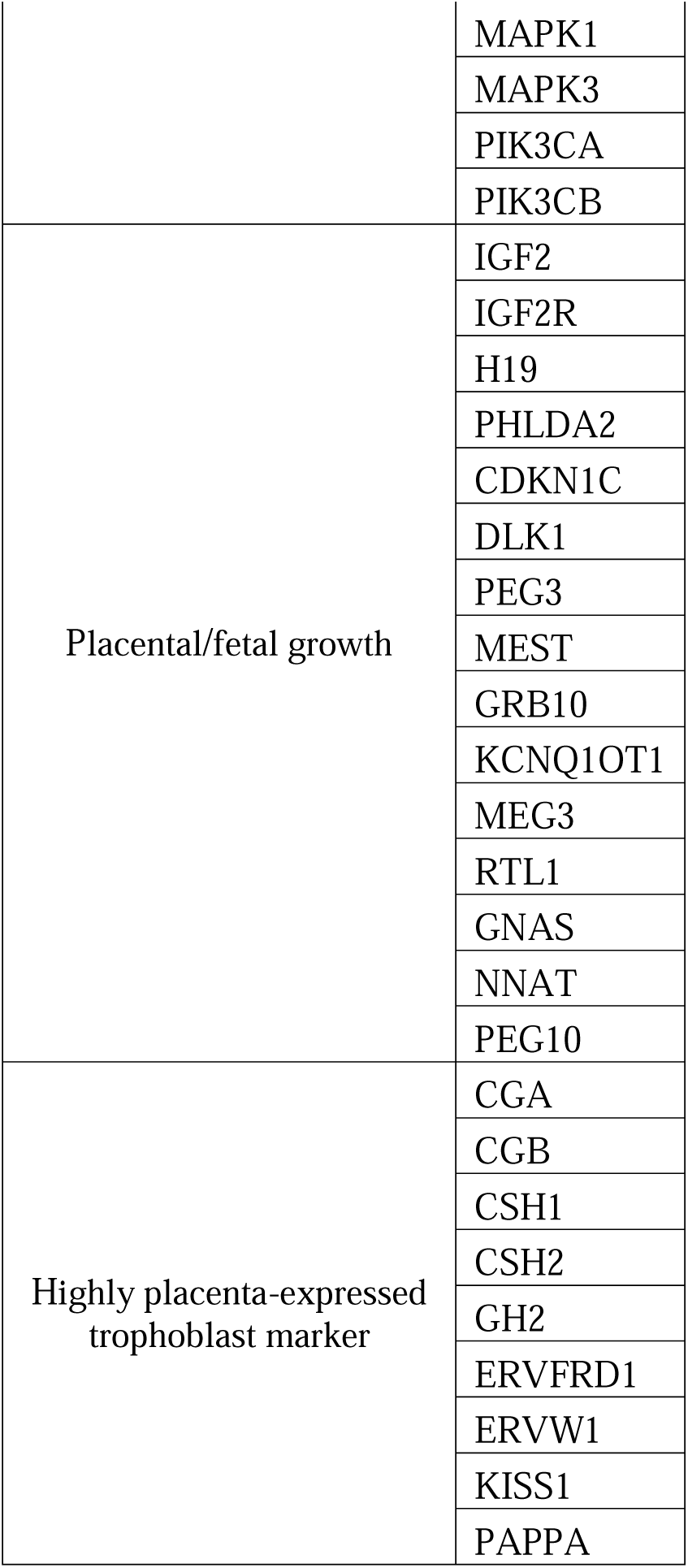
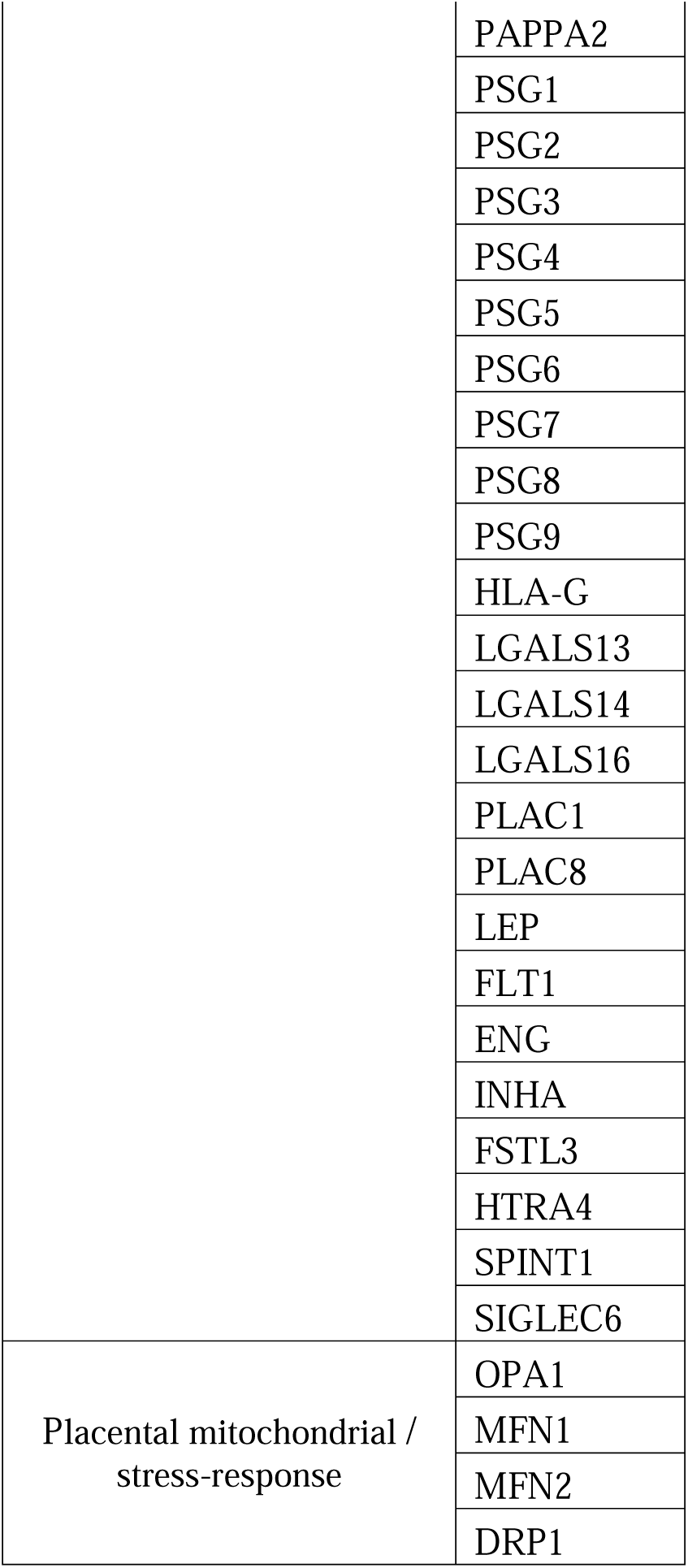
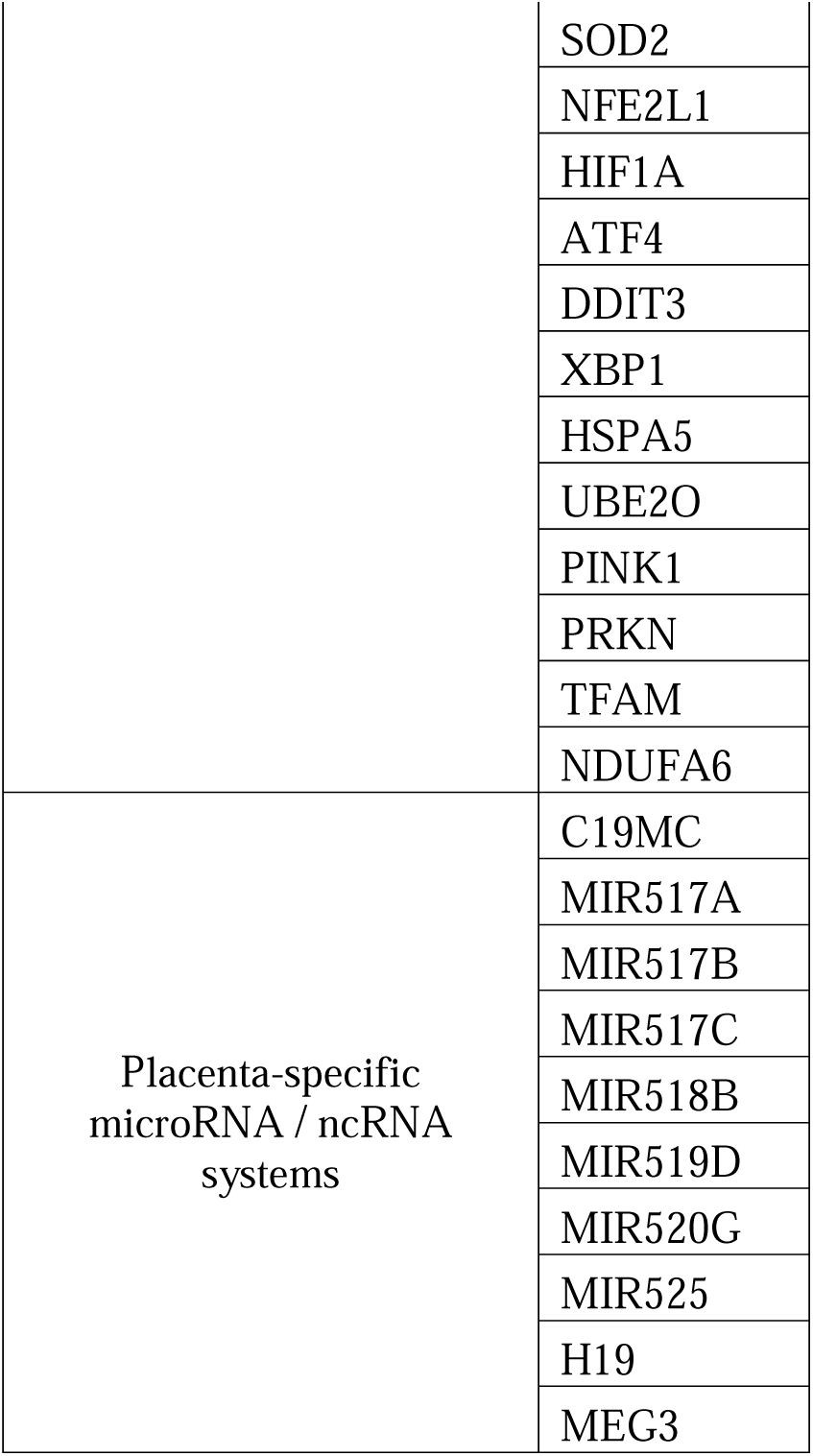
Known placental function candidate genes.

**Table S4.** The observed gene set instances represented in MosaicBase (https://mosaicbase.com/)

| Gene | Category | Number of Instances |
| --- | --- | --- |
| CTNND2 | Any Placental Vascular Malperfusion | 24 |
| PHACTR1 | Any Placental Vascular Malperfusion | 11 |
| SLIT1 | Any Placental Vascular Malperfusion | 2 |
| OPA1 | Any Placental Vascular Malperfusion | 6 |
| IGF2R | Any Placental Vascular Malperfusion | 1 |
| VEGFA | Clinical Placental Insufficiency | 0 |
| SLIT1 | Clinical Placental Insufficiency | 2 |
| DLK1 | Clinical Placental Insufficiency | 1 |
| TEK | Clinical Placental Insufficiency | 4 |
| PHACTR1 | Clinical Placental Insufficiency and/or Any Placental Vascular Malperfusion | 11 |
| GNAS | Clinical Placental Insufficiency and/or Any Placental Vascular Malperfusion | 13 |
| OPA1 | Clinical Placental Insufficiency and/or Any Placental Vascular Malperfusion | 6 |

